# Creatine Kinase is Associated with Bleeding after Myocardial Infarction

**DOI:** 10.1101/19012039

**Authors:** Lizzy M. Brewster, Jim D. Fernand

## Abstract

**Background:** Highly elevated plasma activity of the ADP scavenging enzyme creatine kinase (CK) might reduce ADP and ADP-dependent platelet activation. Therefore, we studied, whether high CK after myocardial infarction (MI) is associated with bleeding.

**Methods:** Data of the Thrombolysis In Myocardial Infarction Study Group phase II trial on the efficacy of angioplasty following intravenous recombinant tissue-type plasminogen activator (rt-PA), are used to assess whether peak plasma CK (CKmax) is independently associated with adjudicated fatal or non-fatal bleeding (primary) and combined bleeding/all-cause mortality (secondary) in multivariable binomial logistic regression analysis, adjusting for baseline and treatment allocation covariates.

**Results:** The included patients (N=3339, 82% men, 88% white, mean age 57 y, SE 0.2), had a history of angina pectoris (55%), hypertension (38%), and/or diabetes mellitus (13%). CKmax ranged between 16 and 55 890 IU/L (mean 2389 IU/L; SE 41), reached within 8 h in 51% of the patients (93% within 24 h). Adjudicated fatal/non-fatal bleeding occurred in 30% of the patients (respectively 26% in the low vs 34% in the high CK tertile), and bleeding/all-cause mortality in 35% (29% in the low, vs 40% in the high CK tertile). The adjusted odds ratio for fatal/non-fatal bleeding (vs not bleeding and survival) was 2.6 [95% CI, 1.8 to 3.7]/log CKmax increase, and 3.1 [2.2 to 4.4] for bleeding/all-cause mortality).

**Conclusion:** Highly elevated plasma CK after MI might be a hitherto overlooked independent indicator of bleeding and hemorrhagic death. This biologically plausible association warrants prospective study of the potential role of CK in hemorrhagic diathesis, and the risk of severe, potentially fatal bleeding with antithrombotic or thrombolytic therapy in the presence of high plasma CK.

**ClinicalTrials.gov identifier (NCT number):** NCT00000505

## Background

Bleeding contributes significantly to morbidity and mortality associated with thrombolytic and antithrombotic drug therapy for ischemic events.^1-5^ With the incidence of major bleeding in acute coronary syndromes (ACS) estimated around 4%, it is well recognized that thrombolytic and antithrombotic therapy involves a fundamental tradeoff between decreasing ischemic risk and increasing bleeding risk. Clinical assessment of bleeding risk is therefore of vital importance.^1-5^

Highly elevated creatine kinase (CK) after myocardial infarction (MI) might reduce ADP-dependent platelet activation in vivo (**Figure 1**).^6^ During myocardial ischemia and infarction, CK and other cell constituents such as myoglobin and troponin enter plasma along with smaller molecules including phosphocreatine.^1-3,6^ This highly elevated CK is proposed to reduce plasma ADP through its scavenging action on ADP, or through conversion into ATP, catalyzing the reaction:

**Figure 1.**
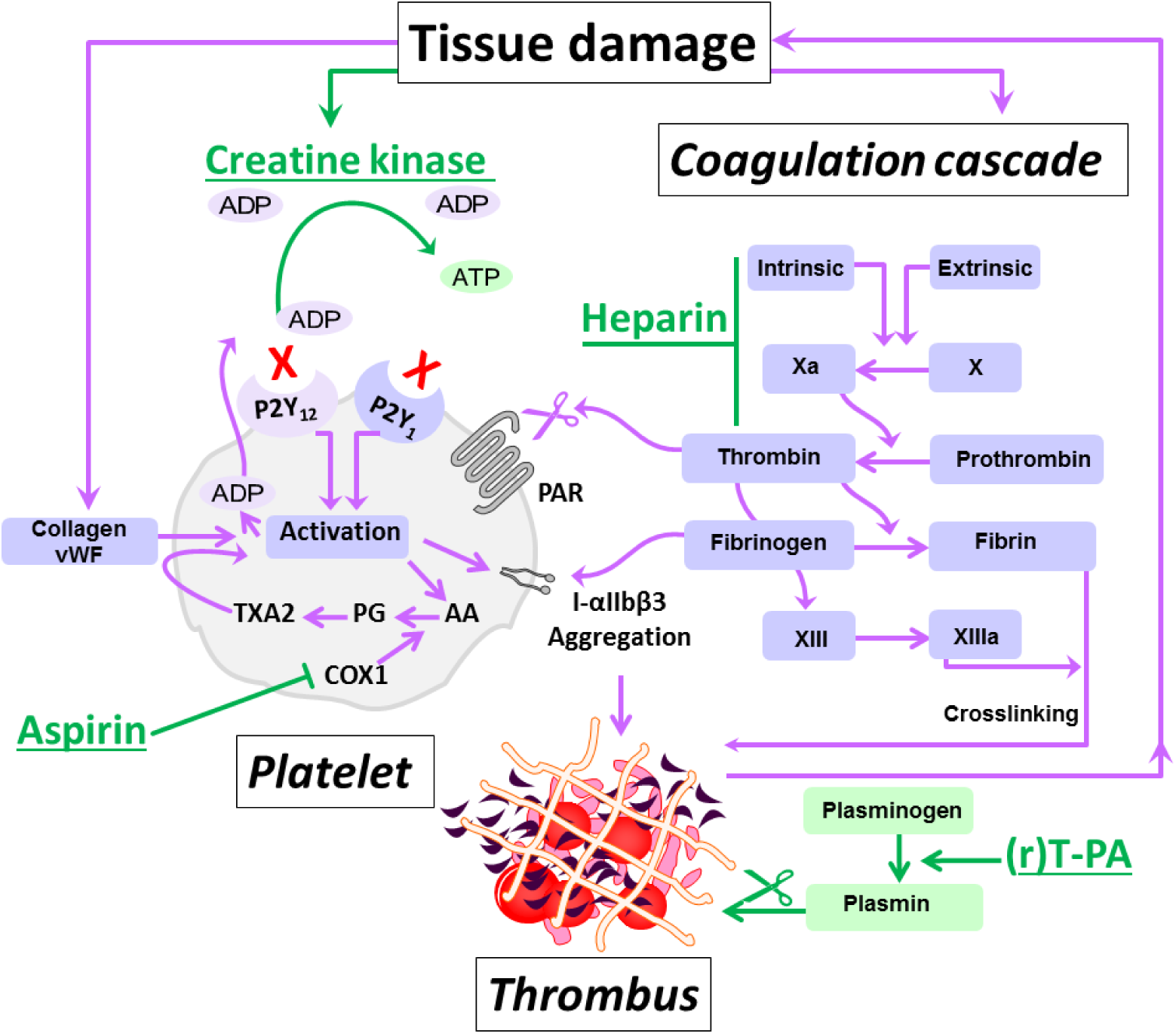
Inhibitory Action of CK on Platelets. Highly schematic representation of thrombus formation and the proposed inhibitory action of the ADP scavenging enzyme creatine kinase (CK) herein. The intrinsic and extrinsic pathways leading to thrombus formation are elaborately described elsewhere.^7-10^ After spontaneous plaque rupture during acute coronary syndrome and during percutaneous coronary intervention, platelets adhere to the injured vessel wall, undergo shape change, cytosolic Ca^2+^ mobilization, and activation (through collagen and von Willebrand factor, vWF). Platelet activation leads to release of adenosine diphosphate (ADP) and thromboxane A2(TXA2) synthesized from arachidonic acid (AA) through prostaglandin (PG) catalyzed by (COX1), which is inhibited by aspirin. These secondary agonists amplify the response to injury and produce sustained platelet aggregation.^7^ In addition, thrombin generated by tissue damage activates platelets’ protease activated receptors (PAR). Platelet activation leads to the generation of more thrombin on the platelets’ surface, which further activates platelets, converts fibrinogen to fibrin, and activates coagulation factors including factor XIII to further stabilize the platelet-fibrin clot. Plasminogen and (recombinant) tissue-type plasminogen activator (r)T-PA bind to the surface of the clot and plasmin degrades fibrin.^7-10^ADP is considered to be central to platelet activation. ADP-stimulation of the P2Y1 receptor activates phospholipase C resulting in weak, transient platelet aggregation. Activation of P2Y12 receptor results in the activation of glycoprotein receptors IIb/IIIa (integrin (I)-αIIbβ3) and firm platelet aggregation.^9^ Creatine kinase has a high binding capacity for ADP, which can be converted to ATP. Highly elevated extracellular CK released during major tissue damage might function to reduce ADP and dampen platelet activation, but might also lead to increased bleeding risk,^6^ in particular when multiple antithrombotic drugs (rT-PA, heparin, and aspirin in TIMI 2) are used.

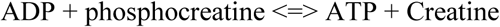

Thus, high plasma CK after MI might increase bleeding risk, in particular when other drugs that affect hemostasis are used. However, once key to the diagnosis of MI, the use of troponin has led to a steady decline in CK estimation after MI and other ACS, and recent data on plasma CK in MI patients are not readily available.^1-4,11^ Therefore, data of the Thrombolysis In Myocardial Infarction phase II trial (TIMI 2)^12-17^ were analyzed to assess the association of CK with bleeding events.

## Methods

### Ethical approval

All patients had given written informed consent to be included in the study.^12-17^ The current analysis was approved by the review board of the Research Ethics Committee of the Radboud University Nijmegen Medical Centre on April 30, 2019 (registration number 2019-5356), and by the US National Heart, Lung, and Blood Institute (RMDA V02 1d20120806) on May 6, 2019.

### The TIMI 2 trial

TIMI 2 was a multicenter trial conducted in 24 clinical centers across the USA. The design and results of the TIMI 2 trial have been described previously.^12-17^ In brief, the study included 3534 patients, all treated with intravenous recombinant tissue plasminogen activator (rt-PA; Genentech, Inc., South San Francisco, California, USA) within 4 hours of the onset of chest pain thought to be caused by ST elevation myocardial infarction (STEMI). Included patients were randomized to either an invasive strategy of cardiac catheterization, and when anatomically appropriate, percutaneous transluminal coronary angioplasty (PTCA) or coronary artery bypass grafting (CABG); vs a conservative strategy, where these procedures were only performed in response to spontaneous recurrent or exercise-induced ischemia.^12-17^ The TIMI 2 trial consisted of 2 major substudies combined into a main study (**Figure 2**). The TIMI 2A study was designed to compare immediate and delayed invasive strategies, and TIMI 2B to compare immediate vs delayed beta-blocker therapy. The main study (n=3339) compared the invasive (n=1681) vs the conservative strategy (n=1658).^12-17^ The primary outcome was non-fatal MI (as defined by CK increase and/or ECG abnormalities) or death within 6 weeks after study entry. Major inclusion criteria were age <76 years, symptoms of ischemic chest pain of >30 minutes duration, and treatment possible within 4 hours of the onset of symptoms. Exclusion criteria included past or present bleeding disorder, significant intestinal bleeding, any recording of blood pressure exceeding 180 mm Hg systolic or 110 mm Hg diastolic during the presenting illness prior to randomization, any history of cerebrovascular disease or transient ischemic attack, oral anticoagulation therapy, advanced illness including malignancies, hepatic, renal disorder, or use of thrombolytic therapy for MI in 2 weeks before admission. During the trial, the initial dose of rt-PA of 150 mg was lowered to 100 mg after the first 520 patients, because of a relatively high number of intracerebral hemorrhages (2.1%), as judged by the Hemorrhagic Event Review Committee (HERC).^18^ Also, the exclusion criteria, such as history of hypertension and history of neurologic disease, were made more stringent.^18^ Adjunctive therapy included intravenous (iv) heparin and aspirin in all patients. Heparin was given as a 5000-unit United States Pharmacopeia (USP) bolus within 1 hour of the start of the rt-PA infusion, followed by an infusion of 1000 units per hour during 4 days guided by the activated partial thromboplastin time. Heparin was given subcutaneously (sc), 10 000 units every 12 hours, from day 5 until hospital discharge. Aspirin dose was initially 80 mg/d starting on the same day as thrombolytic therapy, but in response to the unexpected number of intracranial hemorrhages aspirin therapy was started the day after initiation of thrombolytic therapy. The aspirin dose was increased to 325 mg/d on day 6.

**Figure 2.**
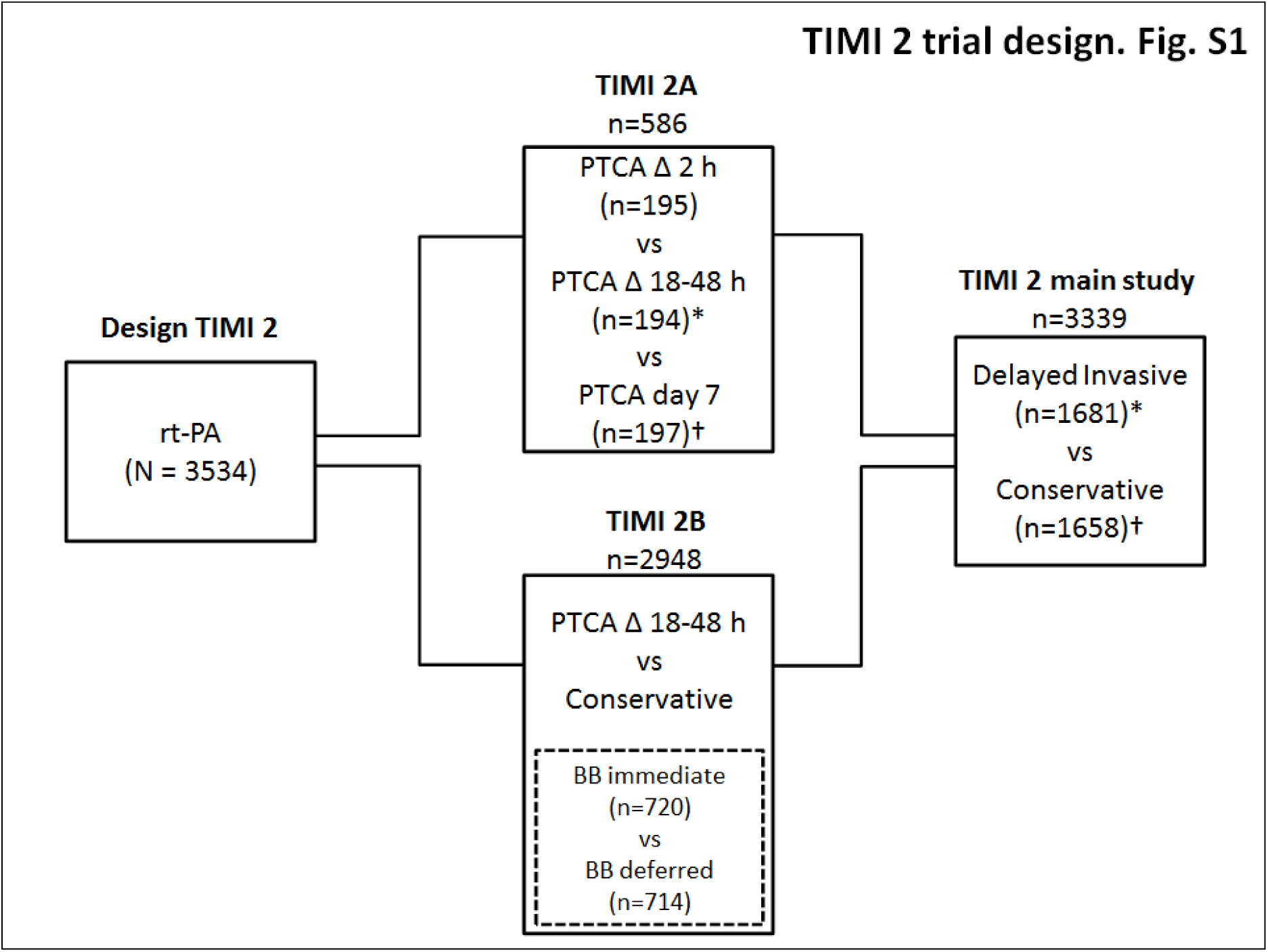
Design of the TIMI 2 trial. Diagram of the Thrombolysis in Myocardial infarction (MI) phase II (TIMI 2) studies. TIMI-2 patients were treated with intravenous recombinant tissue plasminogen activator (rt-PA) within 4 hours of the onset of chest pain thought to be caused by MI, before randomization to an invasive strategy (cardiac catheterization, and when anatomically appropriate, percutaneous transluminal coronary angioplasty (PTCA) or coronary artery bypass grafting within 24 to 48 hours after infarction) or conservatively strategy (with invasive procedures only in response to the occurrence of spontaneous or provoked ischemia). The trial included 3534 patients in 3 subsets: 1) TIMI II main study (large square, n=3339) in which patients were randomized between a delayed invasive strategy (n=1681) and a conservative strategy groups (n=1658); 2) TIMI 2A sub-study n=586), overlapping with the main study but also including an immediate invasive strategy treatment group; and 3) (The beta-blocker substudy (n=1434), a component of main study in which eligible were further randomized between immediate and delayed 3-blocker therapy;^14^ *excluding n=195 immediate invasive treatment; †including participants with PTCA on day 7 (n=197).

### Classification of hemorrhagic events

All bleeding incidents were registered by the local investigator. Furthermore, each patient with blood loss or a reduction in hemoglobin of more than 30 g/L (around 1.9 mmol/L) during hospitalization was reviewed by the HERC. The time of onset and the bleeding site, if known, were registered. Bleeding was classified by the HERC as “major” when a decrease in hemoglobin of more than 50 g/L (around 3.1 mmol/L), intracranial bleeding, or cardiac tamponade was present. Bleeding was classified as “minor” if a hemoglobin reduction >30 and </=50 g/L from an identified bleeding site was present, or if the patient had spontaneous macroscopic hematuria, hemoptysis, or hematemesis. Finally, patients were classified as “loss, no site” if the bleeding site was unknown, but hemoglobin reduction was > 40 g/L (around 2.5 mmol/L) and </=50 g/L. Transfusion of one unit of packed cells or whole blood was counted as 10 g/L when assessing hemoglobin reductions. Bleeding events in trial patients who underwent surgery (mainly CABG) were considered separately.^12-17^

### Classification of the cause of death

The Mortality and Morbidity Classification Committee classified all causes of death. Patients classified as having died from hemorrhage were included under “major hemorrhagic events.”^17^

### Hemostatic Variables

Hemostatic variables (fibrinogen levels, fibrin(ogen) degradation products, rt-PA levels, and plasminogen) were collected on as many patients as possible at baseline, and at 50, 300, and 480 minutes after rt-PA infusion, and determined in a central Coagulation Core Laboratory, using standardized methods as described previously.^12-17^ Platelet count was determined locally at the participating centers.

### CK estimations

Total CK was estimated locally at the study centers, routinely over the first ten days, at baseline and 4 hour (h) intervals during the first day, at 6 h intervals during the second day, and daily at day 3 through 10. The upper reference limit (URL) of each center was recorded.

### Main outcomes

The main outcomes of this analysis are the independent association between peak CK activity (CKmax) with adjudicated fatal or non-fatal bleeding during hospitalization (primary), and with adjudicated combined non-fatal bleeding and all-cause mortality (ACM) (secondary). The tertiary outcome was the association of CK with all investigator reported bleeding.

### Sample size calculation

The probability P of events at the mean value of the CK (and other variables) was conservatively estimated to be 0.25. Sample size was estimated for two levels of the odds ratio (OR) of disease, corresponding to an increase of one standard deviation from the mean value of CK, given the mean values of the remaining variables; at OR 1.2, with at least 1138 participants needed; or at OR 2.0, with at least 95 needed for this analysis with a one-tailed alpha of 5% and 1 – beta of 80.^19^

### Data analyses

Demographic characteristics were analyzed by bleeding status, and the distribution of CK during hospitalization was assessed.^12-18^ Parametric versus nonparametric statistical methods were used where appropriate. However, as extreme CK values are relevant in this analysis, all values were included, with the mean and standard error to the mean (SE) presented, and log transformation to the base of 10 used in regression analysis.^6^ The highest CK activity (IU/L) during hospitalization in an individual patient was used (CKmax), standardized to account for the differences in URL, expressed as a normalized ratio ≥1. In addition, time to CKmax, and time to, severity, and location of the first adjudicated bleeding were assessed. Bleeding events were depicted by CK tertiles, and bleeding severity by CKmax, with multivariable logistic regression analysis to assess whether log CKmax was independently associated with adjudicated fatal or non-fatal severe bleeding (primary), combined (severe) bleeding and all-cause mortality (ACM) (secondary), and all investigator-reported bleeding events (tertiary), compared to no bleeding. Variables were chosen based on previous clinical data,^12-17^ building models adjusting for sex, ancestry (white vs non-white, as defined by the study), age, body mass index (BMI, body weight in kg divided by the squared height in meters), hypertension history, treatment assignment (conservative vs invasive), rt-PA dose, platelet count, and a measure of hemostasis (chosen with Kendall’s tau-b correlation coefficient with adjudicated fatal or non-fatal bleeding), using forced entry and bootstrapping for internal validation. Associations were also modeled excluding bleeding at puncture sites. Missing data are not imputed in the primary analysis, but depending on the number of missing data (>5%), the mechanism (ad random or not), and the data distribution, data will be considered for imputation, with the imputation strategy depending on the missing data analysis. P-values are not reported in this study.^20^ Instead, formal statistical testing is limited and outcomes described and depicted for inspection. Furthermore, the statistical uncertainty surrounding the estimates is communicated, where relevant through 95% confidence intervals (CI) given between square brackets.^20^ Uncertainty regarding the external validity is discussed. Data in parentheses are SE unless indicated otherwise. Statistical analyses were performed with the SPSS statistical software package for Windows version 25.0 (SPSS Inc, Chicago, Ill, USA).

## Results

### Baseline Characteristics and Assigned Treatment

Baseline characteristics of the participants are depicted in **Table 1**. Patients were predominantly white (88.3%), and men (82.1%), with a mean age of 56.8 (SE 0.2) y. Patients had been randomized to the invasive strategy (n=1681) or the conservative strategy (n=1658). Median duration of hospitalization was 10 days, with an interquartile range (IQR) of 8 to 12 days. During hospitalization, all 3339 patients received rt-PA (mean dose 107 (SE 0.3; median 100 IQR), and 98.7% were given heparin, median iv dose for the first 5 days 21 323 U (IQR 17 383 to 25 384); and 19 540 sc for day 6 to 10 (16820 to 21 256). Furthermore, 3201 (95.9%) received antiplatelet agents (99.4% aspirine), 77.5% as monotherapy (99.2% aspirin), 21.3% dual therapy (DAPT; 95.8% aspirine-dypiridamole), and 1.2% triple therapy. Median aspirin dose (n=3181) was 80 mg/day during the first 5 days (IQR 80 to 141 mg) and 325 from day 6 to 10 (275 to 325 mg).

**Table 1.** Patients’ Characteristics

|  | All Participants | No Bleeding | Adj. Bleeding |
| --- | --- | --- | --- |
| N | 3339 | 1389 | 985 |
| Age (y) | 57 (0.2) | 55 (0.3) | 59 (0.3) |
| Men (%) | 82 | 86 | 77 |
| Ancestry (white,%) | 88 | 88 | 89 |
| BMI (kg/m<sup>2</sup>) | 28 (0.1) | 28 (0.1) | 27 (0.1) |
| History of AP (%) | 56 | 51 | 61 |
| History of hypertension (%) | 38 | 35 | 40 |
| History of diabetes mellitus (%) | 13 | 13 | 15 |
| Invasive treatment arm (%) | 50 | 45 | 60 |
| Platelet count ($\cdot 10^9/L$)* | 285 (1.8) | 280 (2.1) | 286 (2.6) |
| IV rt-PA dose (mg/first 6h) | 107 (0.3) | 106 (0.5) | 107 (0.6) |
| P rt-PA 50 min (nanogram/mL)† | 1745 (28.9) | 1667 (42.8) | 1850 (48.7) |
| Plasminogen 50 min (NP %‡ | 68 (0.4) | 70 (0.7) | 65 (0.8) |
| Fibrinogen 8 h (mg/dl)‡ | 185 (1.9) | 195 (2.9) | 173 (3.4) |
| FDP 8 h (microgram/ml)‡ | 292 (16.7) | 233 (23.1) | 400 (39.4) |
| Heparin dose (U/d)§ | 20 532 (98.6) | 21 107 (145.0) | 19 895 (202.0) |
| Aspirin dose (mg/d)§ | 217 (2.8) | 211 (3.1) | 230 (6.6) |
| CKmax (times URL)\|\| | 13 (0.2) | 12 (0.3) | 16 (0.6) |
| Maximum CK (IU/L)\|\| | 55 890 | 20 000 | 55 890 |

**Legend.** Patients by adjudicated, severe non-surgery related fatal or non-fatal bleeding events (primary outcome) vs no bleeding. Remaining patients (n=965), had reported, but non-severe (adjudicated, adj.) bleeding. Data are rounded means (standard errors), unless indicated otherwise. BMI, body mass index, AP, angina pectoris; IV, intravenous; (P) rt-PA, (Plasma) recombinant tissue-type plasminogen activator; NP, % of normal pool; FDP, fibrin(ogen) degradation products; CKmax, maximum creatine kinase (CK) level per patient (mean), expressed as times the upper reference limit (URL, n=3327); Maximum CK, highest CK measured per group. \*At baseline, n=2860; †At 50 minutes after treatment initiation, n=1660; ‡at 8 h after treatment initiation, Plasminogen, n=1484; Fibrinogen n=1695, FDP, n=1670; §mean dose during hospitalization; ||CK peak value

### Main Trial Outcomes

The main findings of the TIMI trial, including recurrent infarction, cardiac function, and survival have been reported previously.^12-18^ PTCA after rt-PA was attempted in respectively 54.0 vs 13.3% of the patients assigned to the invasive strategy vs the conservative strategy. There was no significant difference between treatment arms in the trial’s main outcome of non-fatal MI or death within 6 weeks after study entry, which occurred in 10.9% percent of the invasive strategy group vs 9.7% in the conservative strategy group. In addition, there was no difference in mortality between the immediate vs the deferred beta-blocker therapy groups.

### CK and Bleeding

CK was estimated at 19 different time points, in total 46 523 times in 3327 patients during the hospital stay, from admission (T = 0 h) to day 10 (T = 240 h). Plasma CK ranged from <10 to 55 890 IU/L. Mean CK (SE) per patient was 900 IU/L (SE 14). CKmax ranged between 16 and 55 890 IU/L (mean 2389 SE 41), a mean of 13.5 times the URL (SE 0.2) (**Figure 3, Panel A and B**). Most patients (51.5%) had a peak in CK activity within 8 h (**Figure 3. Panel C**).

**Figure 3.**
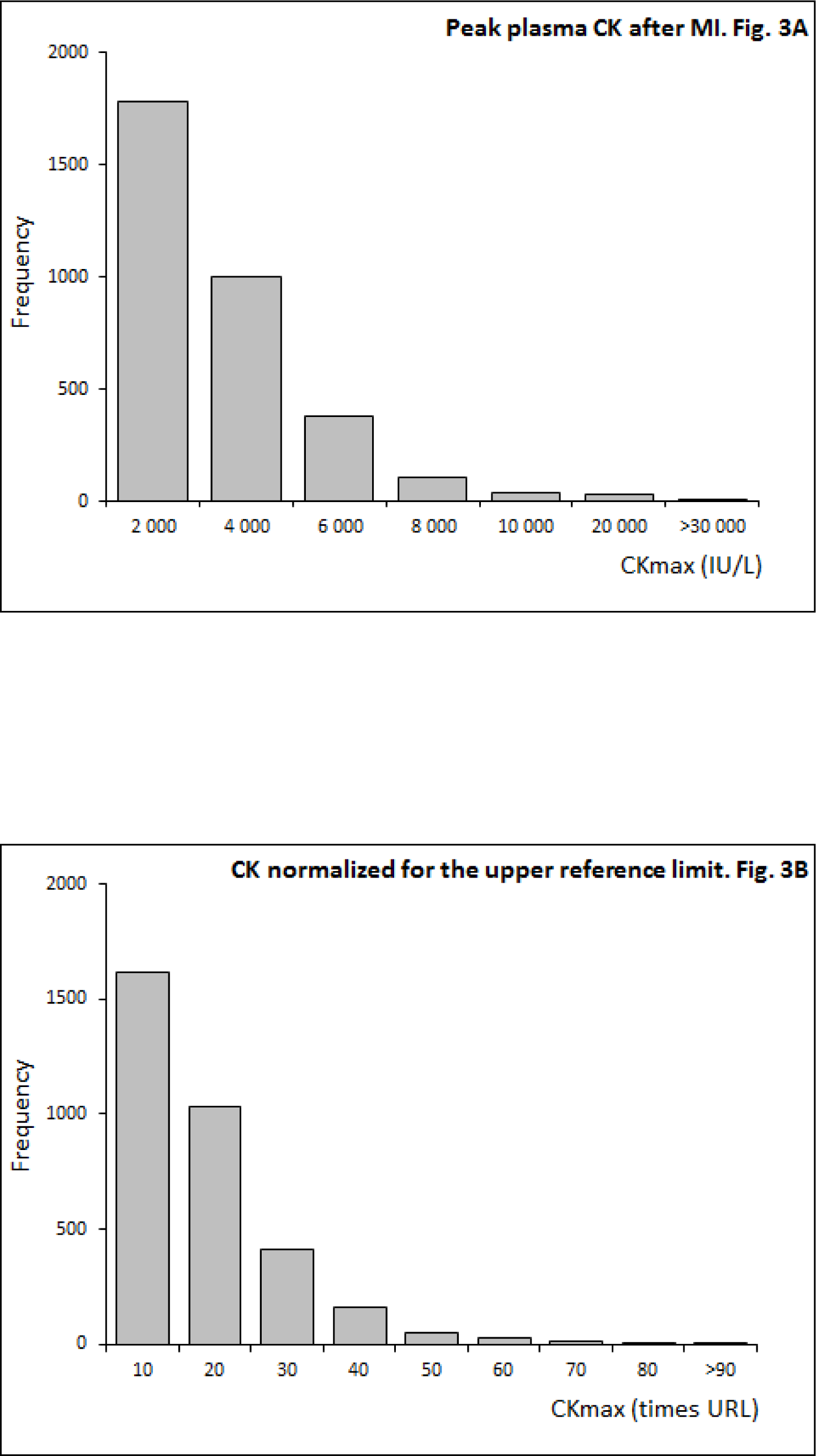
The Association between CK and Bleeding during Hospitalization for MI Panel E-H. Site and severity of bleeding events in relation to CK.

**Figure 3.**
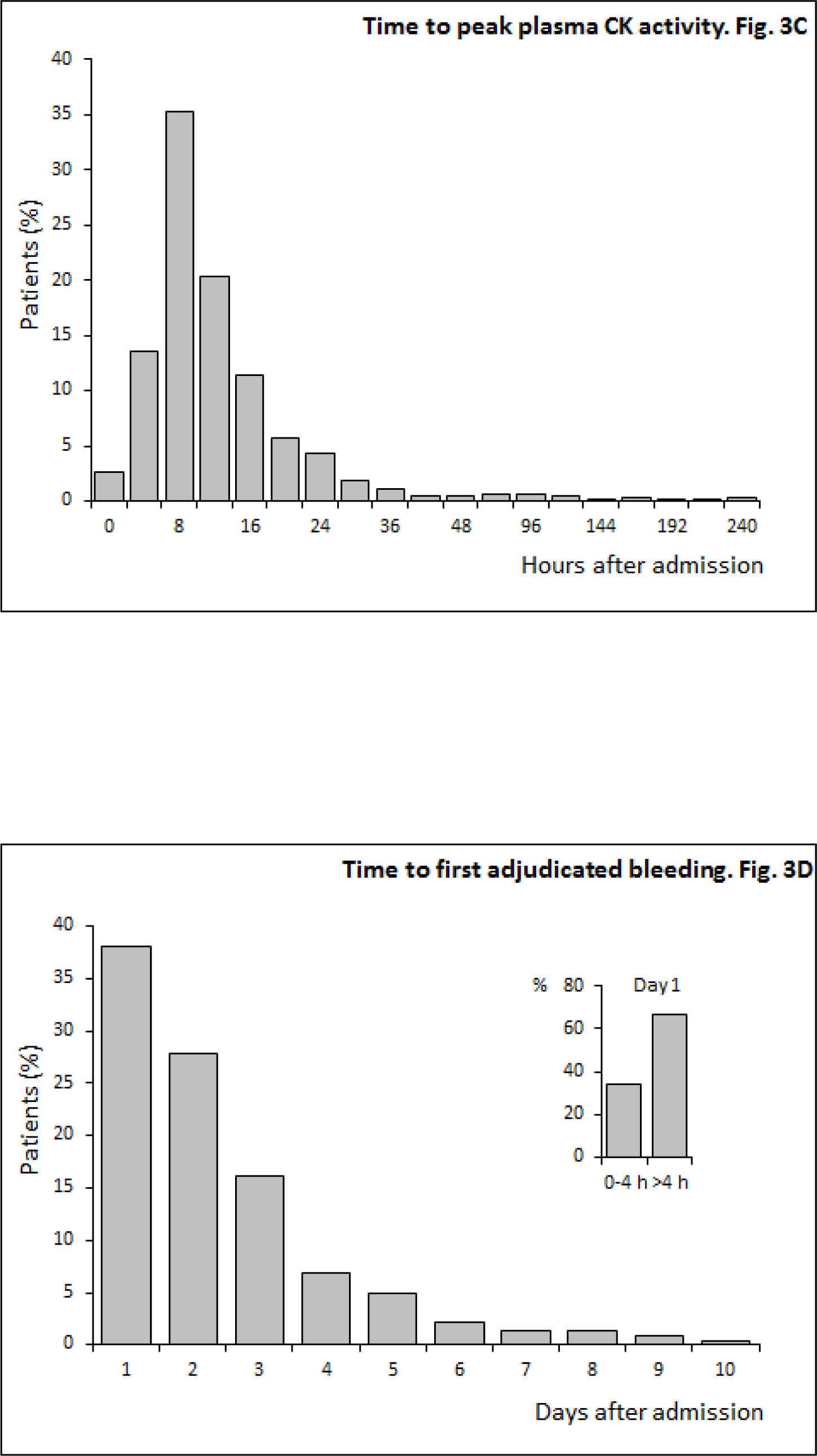
The Association between CK and Bleeding during Hospitalization for MI Panel C-D. Timing of peak CK and bleeding.

**Figure 3.**
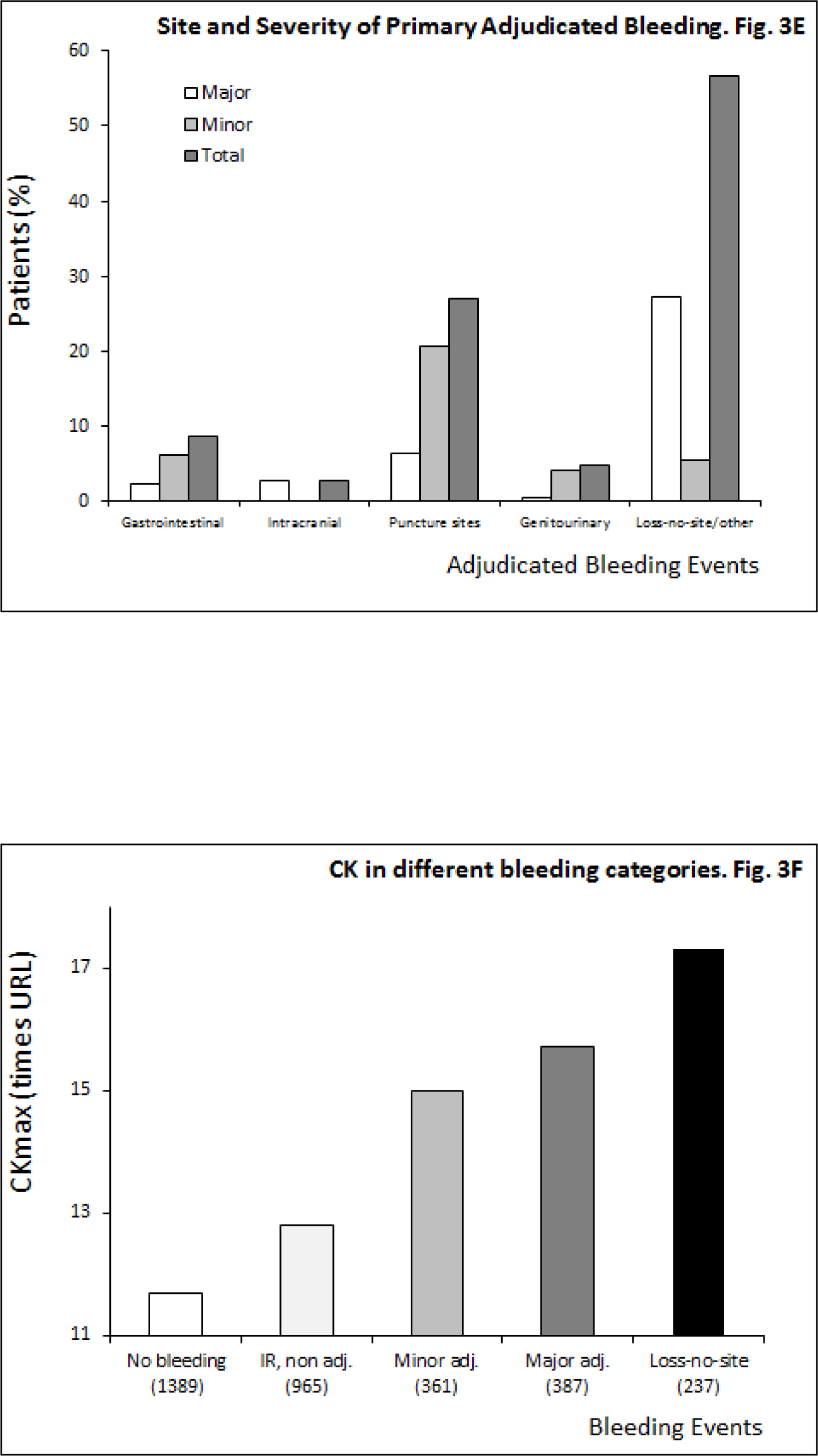
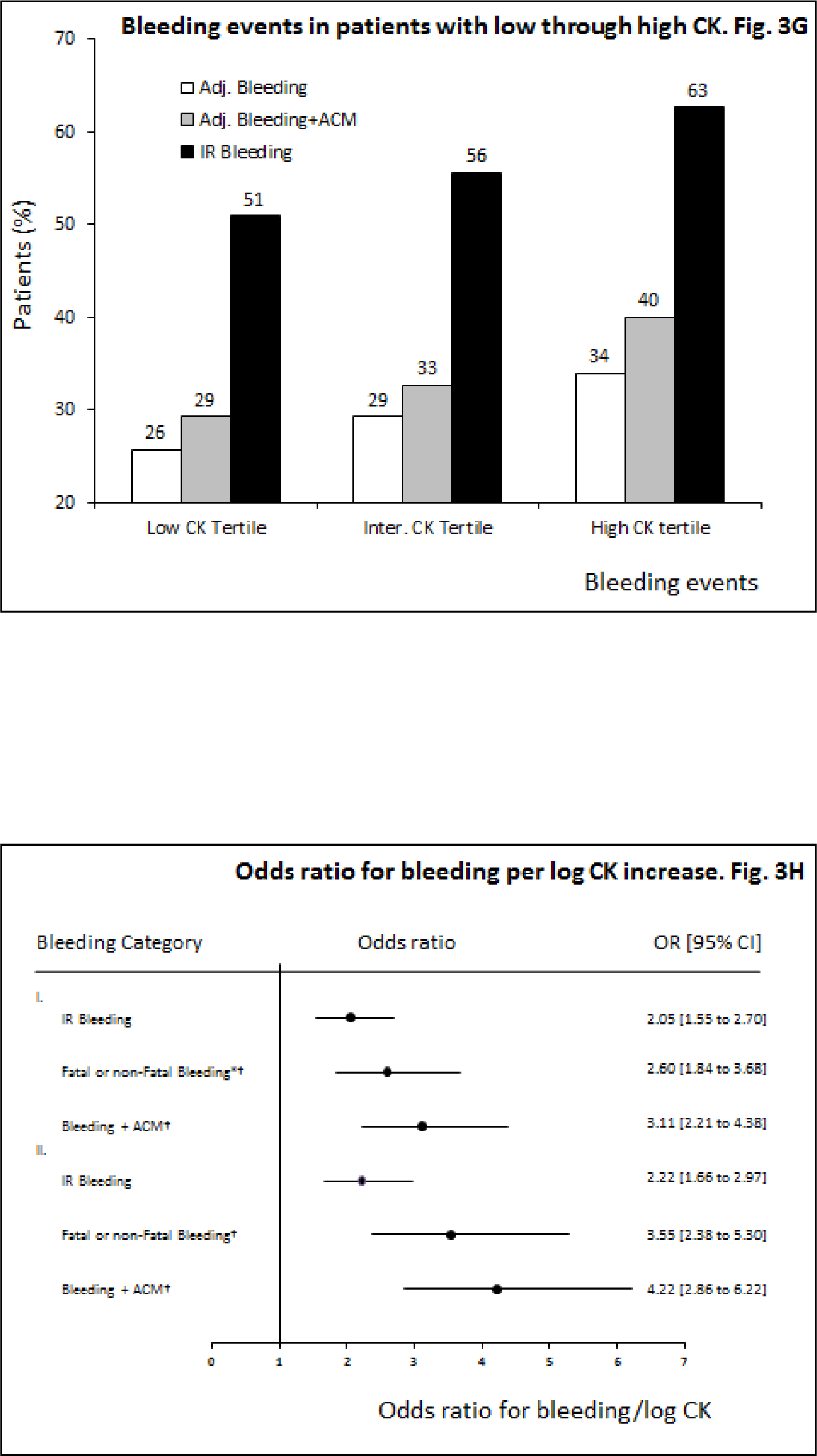
The Association between CK and Bleeding during Hospitalization for MI Panel A-B. Peak plasma CK activity. Panel A to H concern the association of peak plasma creatine kinase (CKmax) with bleeding events. Panel A (IU/L) and B (CK normalized for the upper reference limit, URL), show highly elevated CK activity after myocardial infarction, to more than 90 times the URL. This high CK is thought to inhibit ADP-dependent platelet activation. Panel C and D explores the time to peak CK value (at 8 hours) and time to the first adjudicated bleeding (peak at Day1) Panel E considers the distribution of primary adjudicated bleeding sites by severity, with a relative high frequency of minor bleeding at puncture sites Panel F and G indicate the dose-effect relationship of CK with bleeding. Panel H shows the adjusted odds ratio (OR) with 95% confidence intervals (CI) for bleeding vs non bleeding per log CKmax increase in multivariable binomial regression analysis. I. All bleeding; II. Excluding puncture bleeding. *Primary outcome; †adjudicated bleeding events. Adj. bleeding. First adjudicated bleeding event (fatal or non-fatal bleeding); ACM, adjudicated all-cause mortality. IR bleeding, investigator reported hemorrhagic complication. N=3339; with 12 missing values on CK. Panel H shows results on the subgroup of complete cases without imputation (n = 911 to 1428).

Any bleeding was reported in 1950 patients (58.4%). Of these, bleeding localisations were available in 1878, of whom 705 (37.5%) were registered to have puncture bleedings; 443 (23.6%) hematuria, and 181 (9.6%) gastrointestinal bleedings, as main primary bleeding locations. Non-surgical bleeding events as adjudicated by the HERC (first bleeding site) occurred in 985 (29.5%) of the patients (**Table 1**), 39.3% major bleeding, 36.6% minor bleeding, and 24.1% loss-no-site. Data on the time of bleeding were available in 921 of these patients (93.5%) Adjudicated bleeding occurred on the first 2 days in 57% of the patients (**Figure 3, Panel D**). There was a relatively high occurrence of adjudicated bleeding at vascular sites of catheter placement, venipuncture, arterial puncture, or other vascular instrumentation (n=266; 27.0%; **Figure 3, Panel E**). Blood transfusion was given to 357 patients (36.2%) with adjudicated bleeding events. Eighteen patients (0.5%) died because of bleeding, with a total all-cause mortality of 293 patients (8.8%). It was previously reported that bleeding was associated with different factors including rt-PA dose, hemostasis variables, invasive procedures, age, female sex, low body weight, and a history of hypertension.^12-18^ The timing of beta-blocker therapy had no apparent effect on bleeding risk.^12-18^ The association of bleeding with CK had not been studied previously.

CK by clinical bleeding categories is depicted in **Figure 3, Panel F and G**. The proportion of patients with bleeding events in the low vs the high CK tertile was respectively 26 vs 34% for adjudicated fatal or non-fatal bleeding, 29 vs 40% for combined adjudicated bleeding and ACM, and 51 vs 63% for all investigator reported bleeding (**Figure 3, Panel G**). Finally, multivariable binary logistic regression analysis suggested an independent association between CK and bleeding outcomes (**Figure 3, Panel H**), with a two to four-fold increase in odds for bleeding (and ACM) compared to non-bleeding (and survival) per log CKmax increase, when holding the other variables constant.

## Discussion

### Summary of the Findings

The presented data indicate, to our knowledge for the first time, that plasma CK after myocardial infarction is independently associated with a greater than 100% increase in the odds of fatal or non-fatal bleeding per log CKmax increase (compared to non-bleeding), when conjunctive therapy with antithrombotic and thrombolytic drugs is given. With ample experimental evidence present, the biological plausibility of the association is lined out in Figure 1, suggesting that a massive leak of CK towards the extracellular space and circulation reduces ADP-dependent platelet activation through its binding capacity for ADP, or through conversion into ATP.^6-10^ The data further suggest a temporal and a dose-effect relationship, depicted in Figure 3. The association is potentially clinically relevant. However, inferences from these data are limited. First, the study was not designed to proof that CK causes bleeding. Second, there is a lack of diversity in the patients, who were mainly older, white, men, with one specific condition, MI. Patients with severe hypertension, a condition known to be associated with high CK as well as with cerebral bleeding, were excluded from this trial. Thus, the strength of the association might be over or underestimated in this analysis. Finally, the data need to be considered with caution, as bleeding was typically multifactorial. Patients with higher CK levels probably had larger infarct sizes, and were more likely to be in a poor clinical condition, which might have affected their bleeding risk and survival. Nonetheless, the association found between CK and bleeding can be considered to be hypothesis-generating, and the potential role of CK in hemostasis after tissue injury deserves to be addressed.

### CK: a Tissue Factor that Inhibits Platelet Activation?

Tissue and endothelial factors in thrombosis and hemostasis have been extensively reviewed elsewhere (Figure 1).^7-10^ With vascular injury, thrombus formation involves damage to the vascular wall and exposure to sub-endothelial molecules, initiating the coagulation cascade, adhesion of platelets, thrombus formation, and thrombolysis. Importantly, with damage to high-CK tissue such as muscle and brain, large quantities of intracellular CK may be released into the extracellular space and circulation, along with ADP, ATP, and phosphocreatine.^6,21-24^ ADP conversion or scavenging action by CK will potentially reduce ADP-dependent platelet activation, which might serve to reduce excessive thrombus formation. The presented data accord with a possible association between extracellular CK and inhibition of ADP-dependent platelet activation, which may act in synergy with thrombolytic or antithrombotic drugs to increase bleeding risk. Although the association between CK and platelet function or bleeding risk was not assessed, distinct hypoaggregability of platelets or increased bleeding risk were reported after myocardial infarction.^17,25^ In addition, the ill-understood “trauma-induced coagulopathy” may ensue after major trauma of CK-rich tissue (muscle or brain), associated with inhibition of ADP-dependent platelet aggregation and intractable bleeding.^26^ Also, bleeding events occur more frequently in population subgroups with relatively high tissue and plasma CK, such as in persons of (West-)African ancestry.^6,27-29^ When given fibrinolytic therapy for STEMI, this group had a higher risk of moderate or severe bleeding, adjusted OR 1.36 [1.14 to 1.62], with a hazard ratio of 2.83 [2.08 to 3.86] for hemorrhagic death.^30^ Notably, African ancestry patients had a higher bleeding risk with the use of clopidogrel (adjusted hazard ratio 3.78 [1.35 to 10.60]),^31^as well as with dabigatran.^32^ Whilst these data have no bearing on causality, CK might have affected bleeding risk.

Plasma CK circulates, and although the enzyme is thought to be cleared by the liver,^6^ there is no known inhibitor. As a consequence, its ADP binding effect is probably systemic. The conversion to ATP would need phosphocreatine, which is more likely to be present locally at the site of tissue damage. The role of ATP proper in platelet activation is somewhat enigmatic. ATP is reported to have either no direct effect,^21^ only though inhibition of ADP-dependent platelet activation,^21^ or act on the platelet P2X1 receptor as an inhibitor at low, and an activator at high concentrations.^22^ Importantly, vascular endothelium and blood cells are known to contribute to the interconversion of extracellular adenine nucleotides via ecto-ATPase/ADPase, ecto-5’-nucleotidase, and ecto-nucleotide kinase activities.^23,24^ Thus, the effect of highly elevated extracellular and circulating CK on platelet activation is thought to be mainly through ADP reduction, but the role of ATP, and the potential interaction between CK and ectoenzymes that convert ATP and ADP need further study.

### Clinical Implications and Conclusions

Bleeding is a very relevant clinical issue in thrombolytic and antithrombotic therapy for acute ischemic syndromes, and a major cause of morbity and mortality.^1-5^ With intensification of antiplatelet therapy, adding a P2Y12 inhibitor to aspirin monotherapy (DAPT), as well as prolongation of DAPT to prevent ischemia,^2^ it has become of vital importance to identify patients at high risk for bleeding. Plasma CK might aid in this risk stratification.

The observed association between CK and bleeding is proposed to be present regardless of the definition of ACS or the choice of treatment. Since TIMI 2 was conducted, the use of troponin (cTnI and cTnT) altered the definition of myocardial infarction.^33^ Although currently not routinely assessed, plasma CK activity may be highly elevated in ACS, including STEMI and non-ST segment myocardial infarction (non-STEMI), or after PCI periprocedural myocardial injury or infarction.^33-35^ While all patients in TIMI 2 received rt-PA, primary percutaneous coronary intervention (PCI) is now preferred,^3^ but the association could be suspected in any patient with high CK, whether because of tissue trauma,^26^ (West-)African ancestry,^6^ or use of drugs that increase CK (such as statins), and in particular with the concurrent use of drugs that prevent thrombosis or lyse the clot.^17,30-32^ CK may be the elephant in the room in these conditions, apparently overlooked as a major ADP scavenger molecule in the extracellular space and the circulation. High plasma CK could be relevant in every clinical context of bleeding.

However, more data are needed to substantiate this potential role of circulating CK. Does CK proper inhibit platelet activation in vivo, and how high should CK be to increase bleeding risk? Is such effect local or systemic, how is CK inhibited, and what is the effect of ATP, if any? Is there an interaction with antithrombotic and thrombolytic medication, is plasma CK clinically useful to stratify bleeding risk, and will dose reduction of antithrombotic and thrombolytic drugs reduce the bleeding events and death associated with high CK? These important questions need answers. It is hoped that presenting the theory and these associative data will revive the estimation of CK in patients with ACS to study the association with bleeding, in particular when antithrombotic or thrombolytic drugs are used.

## Data Availability

https://biolincc.nhlbi.nih.gov/

## Acknowledgements

The Manuscript was prepared using TIMI 2 Research Materials obtained from the NHLBI Biologic Specimen and Data Repository Information Coordinating Center. The content does not necessarily reflect the opinions or views of the TIMI 2 study members or the NHLBI.

## Competing Interests

LMB is an inventor on patent WO/2012/138226 (filed). JDF declares no competing interests.

## Sources of Funding

This work is funded by the Dutch CK Science Foundation (national foundation registration number 7106614).

## Notes

### Clinical Trial

NCT00000505

### Funding Statement

No external funding

